# Importation, circulation, and emergence of variants of SARS-CoV-2 in the South Indian State of Karnataka

**DOI:** 10.1101/2021.03.17.21253810

**Authors:** Chitra Pattabiraman, Pramada Prasad, Anson K George, Darshan Sreenivas, Risha Rasheed, Nakka Vijay Kiran Reddy, Anita Desai, Ravi Vasanthapuram

## Abstract

As the pandemic of COVID-19 caused by the coronavirus SARS-CoV-2 continues, the selection of genomic variants which can influence how the pandemic progresses is of growing concern. Of particular concern, are those variants that carry mutations/amino acid changes conferring higher transmission, more severe disease, re-infection, and immune escape. These can broadly be classified as Variants of Concern (VOCs). VOCs have been reported from several parts of the world- UK (lineage B.1.1.7), South Africa (lineage B.1.351) and, Brazil (lineage P.1/B.1.1.28). The conditions that contribute to the emergence of VOCs are not well understood. International travel remains an important means of spread. To track importation, spread, and the emergence of variants locally; we sequenced whole genomes of SARS-CoV-2 from international travellers (n=75) entering Karnataka, a state in South India, between Dec 22, 2020- Jan 31, 2021, and from positive cases in the city of Bengaluru (n=108), between Nov 22, 2020- Jan 22, 2021. The resulting 176 SARS-CoV-2 genomes could be classified into 34 lineages, that were either imported (73/176) or circulating (103/176) in this time period. The lineage B.1.1.7 (a.k.a the UK variant) was the major lineage imported into the state (24/73, 32.9%), followed by B.1.36 (20/73, 27.4%) and B.1 (14/73, 19.2%). We identified B.1.36 (45/103; 43.7%), B.1 (26/103; 25.2%), B.1.1.74 (5/103; 4.9%) and B.1.468 (4/103; 3.9%) as the major variants circulating in Bengaluru city. A distinct clade within the B.1.36 lineage was associated with a local outbreak. Analysis of the complete genomes predicted multiple amino acid replacements in the Spike protein. In total, we identified nine amino acid changes (singly or in pairs) in the Receptor Binding Domain of the Spike protein. Of these, the amino acid replacement N440K was found in 37/65 (56.92%) sequences in the B.1.36 lineage. The E484K amino acid change which is present in both VOCs, B.1.351 and P.1/B.1.1.28, was found in a single circulating virus in the B.1.36 lineage. This study highlights the introduction of VOCs by travel and the local circulation of viruses with amino acid replacements in the Spike protein. These were spread across lineages, suggesting that multiple paths can lead to the emergence of VOCs, this, in turn, highlights the need to sequence and limit outbreaks of SARS-CoV-2 locally. Our data support the use of concentrated and continued genomic surveillance of SARS-CoV-2 to direct public health measures, suggest revisions to vaccines, and serve as an early warning system to prepare for a surge in COVID-19 cases.

## INTRODUCTION

The COVID-19 pandemic caused by the coronavirus SARS-CoV-2 has claimed millions of lives and has affected people living in all parts of the globe^1^. The evolution of the virus did not initially alarm public health specialists or those involved in vaccine development ^2^. However, the emergence of variants with distinct biological properties which include one or more mutations that confer higher infectivity, increased transmission, severe disease, re-infection, and immune escape are a cause for concern^3–9^. Such variants may influence the trend of the pandemic and are therefore broadly known as Variants of Concern (VOCs)^3–8^.

In India, the COVID-19 pandemic began with the importation of the virus in January 2020^10^. It is only after 11 million cases and over 150k deaths that the numbers declined, signalling the end of the first wave of SARS-CoV-2 in the country^1,10,11^. As with other countries in the world, India too started vaccination campaigns in January 2021, at about the same time that reports of VOCs were communicated from the UK, Brazil, and South Africa^3,4,6,11^. The primary concern is that they may herald the second wave of SARS-CoV-2 in the county and/or undermine the vaccination drive.

Genomic studies in India have shown that several lineages of SARS-CoV-2 have been introduced, have spread, and fallen below the limit of detection since January 2020^12,13,22,14–21^. We have previously performed detailed genomic epidemiology of SARS-CoV-2 in the South Indian state of Karnataka, with a population of 64.1 million (Census 2014)^22^. We found multiple introductions of SARS-CoV-2 into the state and at least seven distinct lineages were already circulating in the state by May 2020. Detailed analysis of the contact network of COVID-19 cases to look at transmission within the state emphasized the role of symptomatic individuals in spreading the virus^23^. These data have contributed to our understanding of how the virus enters, spreads, and evolves in a population. In the genomic epidemiology study, no particular lineages were associated with disease severity^22^. Studies of sequences from India juxtaposed with sequences from all over the world, suggest that mutations associated with immune escape and re-infection are already circulating in the population^2,24–26^.

Multiple lineages of SARS-CoV-2 have been reported from across the world and in India ^12,13,15–17,19–22,27^. There are two ancestral lineages of SARS-CoV-2 in the PANGO classification system, A and B^28^. While viruses of both lineages are circulating across the world, viruses of lineage B are more widespread and prominent in number. The viruses responsible for the catastrophic outbreak in Italy, in early 2020, with an amino acid change in the spike protein D614G and were classified into lineage B.1^28^. This lineage is now the dominant lineage across the world. Several studies have now shown that viruses in this lineage transmit better, with increased infectivity in cell culture^29–32^.

Viruses of the lineage B.1 have acquired several other amino acid replacements in the Receptor Binding Domain of the Spike protein – specifically in the lineages which have been designated as VOCs, namely -B.1.1.7 (N501Y), B.1.351 (N501Y, E484K, K417T) and P.1 from the lineage B.1.1.28 (N501Y, E484K, K417T). Some of these amino acid replacements either singly or in combination have been shown to influence transmission of the virus, interfere with neutralization of the virus, and are associated with an increase in the number of hospitalizations^2,5,7,8^. The spread of these lineages, therefore, has global implications^5,33^. Early data suggests that some variants may escape neutralization by both therapeutic antibodies and antibodies induced by previous infection and vaccination ^8,9,34,35^. This has implications for the efficacy of Spike sequence-based vaccines and suggests that re-infection is possible^7,36^.

Rapid sharing of genomic information enabled the global community to pick-up cases of VOCs and implement relevant public health measures^3,4,6^. A concentrated, ongoing, local approach to genomic surveillance is critical for the identification of variants and establishing epidemiological links with the trend of the outbreak^5,7,12,22^. This has also proved critical for local outbreak management and informed policy decisions across the world^5,7,37,38^.

It is in this context that we conducted genomic surveillance of COVID-19 positive international travellers to the south Indian state of Karnataka between Dec 22, 2020-Jan 31, 2021 (n=75). We also performed sequencing of SARS-CoV-2 (n=108), collected between Nov 22, 2020-Jan 22, 2021) in Bengaluru city (Bengaluru Urban District) to identify and track locally circulating variants and potential VOCs.

## METHODS

### Samples for Sequencing

The Department of Neurovirology, at the National Institute of Mental Health and Neurosciences (NIMHANS), Bengaluru, is an ICMR (Indian Council of Medical Research) approved COVID-19 diagnostic centre. Further, the Government of Karnataka and the Government of India designated our lab as a nodal centre for genomic sequencing. This study was granted a waiver by the Institutional Ethics Committee of NIMHANS in light of the public health emergency.

Nasopharyngeal and oropharyngeal swabs collected from International Travellers (n=75, Dec 22, 2020 – Jan 31, 2021), samples from COVID-19 cases in Bengaluru city (n=108, Nov 22, 2020-Jan 22, 2021), and from a local outbreak (Feb 2021) were included in the study. Of the 42 samples collected from the local outbreak, 14 were suitable for sequencing (RT PCR positive, Ct value < 30) and were analysed further.

### RNA Extraction and RT-PCR

Nucleic acid extraction was performed with automated magnetic bead-based extraction method, using the Chemagic Viral DNA/RNA special H96 kit (PerkinElmer, CMG-1033-S) following manufacturer’s instruction. SARS-CoV-2 detection was done using ICMR approved diagnostic kits. A total of 197 RT PCR positive samples fulfilling the following criteria – i. Ct values less than 30 in the case of international travellers (n=75), and local outbreak (n=14) or ii. Ct value less than 25 for local cases (n=108), were taken for whole genome sequencing.

### Whole Genome Sequencing of SARS-CoV-2

Whole genome sequencing was performed using the amplicon sequencing approach described in the ARTIC Network protocol using the V3 primer set^39^. The resulting amplicons from 12-24 samples were barcoded using the native barcoding kits (NBD104/114, Oxford Nanopore Technology (ONT)) and sequencing libraries were prepared using the ligation sequencing kit (SQK-LSK109, ONT). The barcoded library was loaded on to FLO-MIN-106 flow cells and sequenced on the MinION (ONT). An average of 0.12 million (median) sequencing reads were acquired per sample with a median coverage of 1737x (Supplementary Table1).

### Analysis of sequencing data and data sharing

Analysis of sequencing reads was performed as described previously^22^. Briefly, sequences were basecalled and demultiplexed using guppy (v3.6). Amplicon sequencing primers were removed from the reads by trimming 25bp at the ends and using BBDuk (v38.37). Reference mapping based assembly of the genomes was performed using Minimap2 ver 2.17 using NC_045512 as the reference. A consensus genome was generated with a coverage cut-off of 10x and the 0% majority rule. This was then edited, and aligned to the reference for annotation. Of the 183 samples from international travellers and local cases, 176 (73/75 imported, 103/108 circulating) genomes could be used for the determination of lineage using the PANGO web application (Pangolin v2.2.2 lineages version 2021-02-12) ^28^. Of the 176 genomes,162 were complete (>92% at 1X and >85% at 10X) and were deposited into the GISAID Database^40^, accession numbers are provided in Supplementary Table 2. Complete sequences (162) were analysed for SNPs and amino acid replacements with reference MN908947.3 (Wuhan-Hu-1) using the CoV-Glue Web Application^41^.

### Phylogenetic analysis

A total of 168 genomes, including the 162 described above, and an additional 6 complete genomes from a local outbreak, were used for phylogenetic analysis with the reference NC_045512 as an outgroup. Multiple sequence alignment was performed using MUSCLE and a maximum likelihood tree was constructed using iqtree^42,43^. The GTR+F+I+G4 substitution model was found to be the best-fit model (of the 88 models tested) using the Bayesian Information Criterion. The consensus tree was constructed from 1000 bootstraps and bootstrap values over 70 were interpreted.

## RESULTS

We sequenced SARS-CoV-2 genomes from 197 SARS-CoV-2 positive individuals, including international travellers (n=75), local cases (n=108), and a local outbreak (n=14). Lineage classification using the PANGO scheme was possible for 176 genomes which were either imported (73/75) or circulating (103/108) (Fig 1 A, B), and for all 14 genomes from the local outbreak (Supplementary Table 3). The genomic surveillance for the local outbreak was carried out to identify the lineage/lineages responsible for the outbreak (Fig 1C).

**Fig 1.**
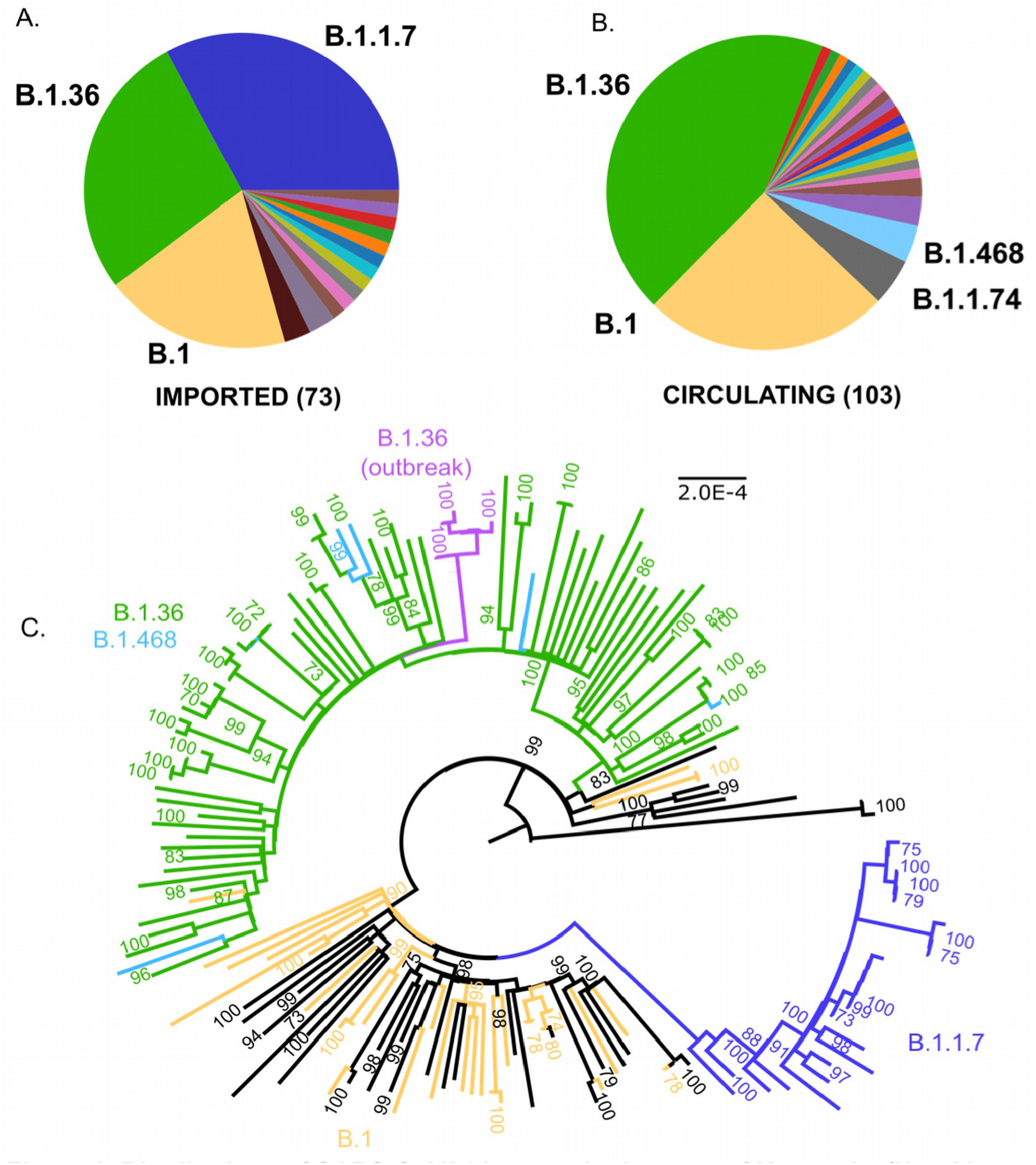
Distribution of SARS-CoV-2 Lineages in the state of Karnataka (Nov 22, 2020- Jan 31, 2021). A. SARS-CoV-2 lineages imported by international travel into Karnataka (n= 73) B. SARS-CoV-2 lineages circulating in Bengaluru city (n= 103). Colours represent different lineages. Lineages with greater > 4 sequences are labelled. C. Maximum Likelihood Phylogenetic tree of 168 complete SARS-CoV-2-genomes from Karnataka, rooted by the reference genome (NC_045512). The scale (length of branches) is in substitutions per nucleotide site. Predominant lineages are coloured. Sequences from a local outbreak of SARS-CoV-2 are coloured in magenta. Numbers on the nodes indicate bootstrap support values (in %), values above 70 are shown.

A total of 34 lineages were detected from the 176 genomes in this study. A complete list of lineages and their frequencies is provided in Supplementary Table 4. Briefly, genomes from imported and circulating viruses belong to both A (3/176) and B (173/176) lineages. Within A, two (2/103) circulating genomes were classified into A.23.1. Of the 173 genomes in lineage B, two genomes were classified into lineage B (2/173), the rest were derived from B.1(130/173) or B.1.1(41/173).

The genomes from imported cases grouped into 16 distinct lineages (Fig 1A, Supplementary Table 4) including B.1.1.7 (24/73, 32.9%), B.1.36 (20/73, 27.4%) and B.1 (14/73, 19.2%). The first introduction of B.1.1.7 was noted in the last week of December 2020, and by January 31, 2021, this lineage made up 32.9% (24/73) of all imported cases (Fig 1 A, Supplementary Table 4).

Circulating genomes grouped into 24 distinct lineages, dominated by the lineages B.1.36 (45/103; 43.7%), B.1 (26/103; 25.2%), B.1.1.74 (5/103; 4.9%) and B.1.468 (4/103; 3.9%) (Fig. 1 B, Supplementary Table 4). Only a single sequence of B.1.1.7 was detected during the study period as part of this surveillance effort in a non-traveller. Sequences from the lineage B.1.36 and derived lineages (70/176) grouped into a distinct phylogenetic clade together with sequences belonging to lineage B.1.468 (6/176) (Fig 1C).

Genomic investigation of an outbreak of SARS-CoV-2 in the city of Bengaluru in early Feb 2021, revealed that 14/14 sequences from the outbreak could be classified into lineage B.1.36. Complete genome sequences could be recovered from 6/14 cases. All six viruses grouped into a clade within the largely B.1.36+B.1.468 clade (Fig 1C).

Of the 176 genomes from travellers and in circulation, for which lineage classification was possible,162 complete genomes (with coverage > 92% at 1X and > 85% at 10X) were used for the analysis of SNPs and amino acid replacements. A total of 968 SNPs (Supplementary Table 5) and 529 amino acid replacements (Supplementary Table 6) were identified. Of these amino acid replacements 61 were in the Spike protein of circulating viruses, and 32 in Spike protein of imported viruses (Supplementary Table 7,8). The B.1.36 lineage had 226 amino acid replacements, 31 of these were in the Spike protein. Although only the D614G and N440K were present in an appreciable number (> 50%) of sequences (Supplementary Table 9).

We carried out further analysis of the amino acid replacements in the RBD domain of the spike protein (Fig 2A, Supplementary Tables 7,8) and mapped them on the Maximum-Likelihood tree (Fig 2B). We identified mutations leading to nine amino acid replacements in the RBD (Fig 2A). Of these, five (S477N, E484K, E484Q, S494L, S494P) were found in viruses circulating in Bengaluru, and the amino acid replacement V483A was from an imported case. The N501Y change was confined to the B.1.1.7 lineage. The N440K replacement was seen in 45/76 (59.2%) sequences in the B.1.36+B.1.468 clade (Fig 2B) and 37/65 sequences in B.1.36 lineage. Of the six sequences from a cluster of cases (Outbreak), only a single sequence carried the mutation resulting in the N440K change (Fig 2B, Supplementary Table 10). A single branch of the B.1.36+B.1.468 clade (n=4, 3 of which were imported) had an additional amino acid replacement F490S in the RBD (Fig 2B). The mutations in the RBD were seen across the phylogenetic tree and clades (Fig 2B).

**Fig 2.**
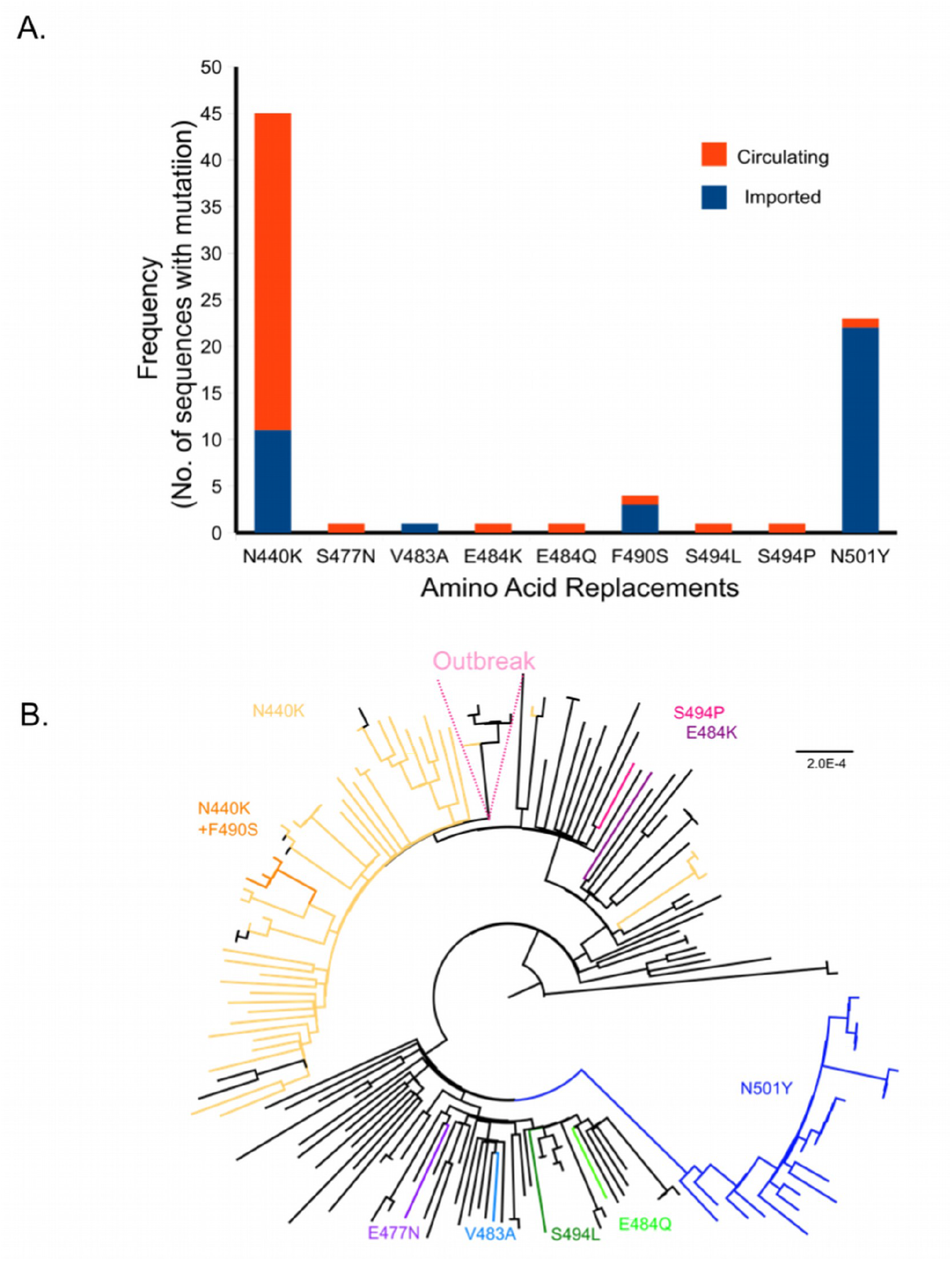
Amino acid replacements in the Receptor Binding Domain (RBD) of SARS-CoV-2. A. Frequency of amino acid replacements in the RBD (amino acids- 387- 516 in the Spike protein) is shown as a bar graph. Frequencies are plotted against the amino acid replacement. Orange and blue represent the circulating and imported genomes respectively. B. Maximum Likelihood phylogenetic tree highlighting branches with the indicated amino acid substitutions.s

## DISCUSSION

In this study, we found 34 lineages of SARS-CoV-2 circulating and imported into Bengaluru city in Karnataka, India, between Nov 22, 2020 – Jan 31, 2021. We aimed to detect the introduction of the global VOCs (lineages B.1.1.7, B.1.351, P.1/B.1.1.28), as well as genotype the variants of SARS-CoV-2, circulating since our last study, which highlighted the introduction and spread of seven lineages of SARS-CoV-2 in Karnataka, between March-May 2020^22^.

We found no evidence suggesting that the B.1.1.7 lineage was present in Karnataka before late-Dec 2020. We first detected the B.1.1.7 variant in Karnataka, in an international traveller from a sample collected on Dec 22, 2020 (Supplementary Table S3). The first and only case of non-travel related B.1.1.7, in our study, was detected in the middle of Jan 2021 in an individual who was in contact with an international traveller(Supplementary Table 3). These data together suggest that B.1.1.7 in Karnataka was limited to travel-associated cases and was not in the community during the study period. At the end of the study period, the B.1.1.7 lineage was detected in 32.9% of all imported cases (Supplementary Table 4). We did not detect the variants P.1/B.1.1.28 or B.1.351 reported from Brazil and South Africa respectively in this study.

We found that B.1.36 and B.1 lineages dominated in both the imported (20/73; 27.4%,14/73, 19.2%) and circulating viruses (45/103; 43.7%, 26/103; 25.2%) in our study (Supplementary Table 4). B.1.36 was first reported from Saudi Arabia in Feb 2020 (Supplementary Table 11) and has now been reported from many parts of the world including India. In our earlier work in Karnataka, we detected only two samples (2/91, 2.2%) clustering into this lineage in the middle of May 2020, which were then classified under the parent lineage B.1. Of the 176 sequences in the present study, 65 sequences were classified into B.1.36 (36.9%) and five were classified as B.1.36 derived lineages (2.8%) (Supplementary Table 4). The B.1.36 lineage was both imported by international travel (20/73) and circulating (45/103) in Bengaluru city (Supplementary Table 4). The lineage is characterized by the following amino acid replacements-nsp12-P323L(95.38%), S-D614G (93.85%), S-N440K (56.92%), ORF 3a-Q57H (90.77%), ORF 3a-E261*(81.54%), nsp3-T183I (81.54%), nsp16-L126F(80%), N-S2P (72.31%), ORF 8-S97I (72.31%) (Supplementary Table 9). The immune escape associated amino acid change, N440K has been reported from the states of Andhra Pradesh, Maharashtra, Telangana, and Karnataka, and is also associated with reinfection^24,36,44^. This change was found in 37/65 (56.92%) of the sequences clustering to B.1.36 (Supplementary Table 9).

An outbreak of SARS-CoV-2 occurred in Bengaluru in early Feb 2021, raising concerns about the spread of variants, the threat of a second wave, and reduction in the efficacy of vaccines. This outbreak in a college where students were returning from different states within India was driven by related viruses belonging to the B.1.36 lineage (Fig 1C, Supplementary Table 3). Only one of the six sequences from the outbreak cluster had the mutation resulting the N440K replacement in the Spike protein (Fig 2C, Supplementary Table 10). This supports the idea that mutations in gene encoding the Spike protein may arise sporadically/multiple times in different clades.

Apart from the introduction and spread of known VOCs, the emergence of variants locally is also a cause for concern. Early in the pandemic, a single mutation in the gene encoding the Spike protein of SARS-CoV-2 resulting in a D614G amino acid change was identified to increase infectivity and transmission^2,29,32^. Viruses with this amino acid replacement dominate across the globe^31,45^. Mutations in the gene encoding the Spike protein are of particular concern due to the role of this protein and its Receptor Binding Domain (RBD) in viral binding and entry^46^. Some of these mutations have been shown to increase infectivity, affinity to the ACE-2 receptor or affect neutralization by antibodies i*n vitro*. Viral genomes with these mutations were already circulating viruses by mid-2020 ^2,25,26,44,47,48^.

In the sequences from this study, nine amino acids replacements were noted in the RBD domain of the Spike protein (Fig 2B, Supplementary 7-8). They occurred singly or in pairs (N440K+F490S) (Fig 2). All nine amino acid changes, namely N440K, S477N, V483A, E484K/Q, F490S, S494L/P, N501Y are associated with immune escape^24,25^. Viruses with some of these amino acid changes were already known to be circulating in other parts of India ^16,17,24^.

Mutations in the gene encoding Spike protein that do not map to the RBD have also been described; particularly near the polybasic cleavage site at the S1/S2 boundary of the Spike protein. Towards the end of the year 2020, multiple lineages with amino acid replacements at position 677 were noted^49^. Four viruses in our study have mutations resulting in amino acid changes at this position (Q677H (n=3), Q677P (n=1)) (Supplementary Table 6).

It is to be noted that in this study we have only included samples with Ct values less than 25 for surveillance of circulating SARS-CoV-2 genomes and Ct values less than 30 for sequencing of international travel-related cases. We have also sequenced only a fraction of cases in a limited geographical area. This may therefore present an incomplete view of circulating viruses and inflate the ones that are more readily sequenced. Also, as we have used the amplicon sequencing approach, not all regions of all lineages are well covered by sequencing reads. Others have also noted homoplasy in SARS-CoV-2, this highlights the need to be cautious while interpreting the phylogenetic relationships between SARS-CoV-2 sequences, especially in the context of outbreaks^50^.

In summary, our data highlight an increase in the frequency of the lineage B.1.36 in Bengaluru Urban, in Karnataka, and importation events indicate an underappreciated global burden (Fig 1, Supplementary Table 4). Whether this increase is because of epidemiological linkages such as increased travel, continued local transmission chains or super-spreader events remains to be determined. It is beyond the scope of this work to examine whether the lineage, contributing mutations, and amino acid changes impact transmission/infectivity of the virus. Our data emphasize that a consolidated and local approach to genomic surveillance which includes sequencing of SARS-CoV-2 from travellers, circulating variants, and outbreaks, in a continuous manner is necessary to detect VOCs. Rapid identification of such variants can aid in preparing the healthcare system for a surge in cases, suggest revisions to vaccines and diagnostic tests, inform the international community, and guide public health measures.

## Data Availability

All sequence data have been deposited in the GISAID database. All other data, including the accession numbers for the sequences, are included in Supplementary Information.

https://www.epicov.org/

## SUPPORTING INFORMATION

Supplementary Table 1-Summary of sequencing results

Supplementary Table 2 – GISAID Accession ID for sequences

Supplementary Table 3 – Details of sequenced samples

Supplementary Table 4 – PANGO lineage assignments for SARS-CoV-2 genomes

Supplementary Table 5 – Position and frequency of single nucleotide polymorphisms

Supplementary Table 6 – Position and frequency of amino acid replacements

Supplementary Table 7-Amino Acid Replacement in Spike Protein (Imported)

Supplementary Table 8 – Amino Acid Replacement in Spike Protein (Circulating)

Supplementary Table 9 – Amino acid Replacements in lineage B.1.36

Supplementary Table 10-Position of amino acid replacements in the Spike protein of sequences from a local outbreak

Supplementary Table 11 – Acknowledgement for sequences from GISAID

## AUTHOR DECLARATION

The authors declare that they do not have any financial or non-financial relationships that could present a conflict of interest.

## ACKNOWLEDGEMENTS

This work would not have been possible without the support of the Government of Karnataka, State Surveillance team for COVID-19, in particular Ms. Prameela Dinesh, Directorate of Health and Family Welfare Services, Government of Karnataka. We would like to thank all the labs and Primary Health Care centres that collected samples for testing and genomic surveillance. We would like to thank the COVID testing lab in NIMHANS. We would also like to acknowledge Prof. Sudhir Krishna’s laboratory at the National Centre for Biological Sciences (NCBS) for support with reagents, NCBS for access to their computational resources for carrying out our analysis, and Dr. Farhat Habib for custom scripts used in data analysis. We gratefully acknowledge the contributions of all the laboratories that have submitted their sequences to GISAID, in particular laboratories across India that have been involved in sequencing efforts.

